# A multi-center study to discern the diurnal variation of wearable device-based heart rate variability (HRV) in the Chronic Renal Insufficiency Cohort (CRIC) Study

**DOI:** 10.1101/2025.04.30.25326177

**Authors:** Carsten Skarke, Wei Yang, Daohang Sha, Nicholas F. Lahens, Tamara Isakova, Mark Unruh, Rajat Deo, Eunice Carmona-Powell, John H. Holmes, Elaine Ficarra, Jing Chen, Jiang He, Hernan Rincon-Choles, Vallabh Shah, Chi-yuan Hsu, Amanda H. Anderson, James P. Lash, Mahboob Rahman, the CRIC Study Investigators

## Abstract

Little is known about the prognostic value of out-of-clinic biometric monitoring of cardiovascular function in chronic kidney disease (CKD). Using real-world sampling strategies, a mean (±SD) of 50.3±9.3 hours of ECG recordings from wearable BioPatch ECG devices was collected in a cohort consisting of 458 participants from seven Chronic Renal Insufficiency Cohort (CRIC) centers. The presence of diabetes was associated with a 7.4 ms lower Standard Deviation of NN Intervals (SDNN) compared to non-diabetic participants (*p*=0.001). Multivariable linear regression revealed that participants without proteinuria (uPCR<0.2) had a 5.15 ms higher SDNN compared to participants with proteinuria (uPCR≥0.2, *p*=0.027). Cosinor modeling suggested differences in SDNN acrophase quartiles for diabetes (*p*=0.02), history of cardiovascular disease (*p*=0.003), eGFR (*p*=0.04), systolic blood pressure (*p*=0.04), and beta blocker use (*p*=0.0002). In the spline analysis, the SDNN curve differed between participants with and without cardiovascular disease (*p*=0.0005). This study assembled the largest dataset to date of SDNN as an index for heart rate variability from wearable digital health technology in the CRIC. The study demonstrates that several clinical and demographic factors are associated with SDNN in participants with CKD. This sets the stage to determine the predictiveness of time-specific HRV metrics for future clinical outcomes.

## Introduction

Kidney and heart function are regulated by the molecular circadian clock to adapt to time-specific changes in physiological needs over the 24-hour cycle to maintain a healthy homeostasis (Juffre and Gumz 2024; Thosar, Butler, and Shea 2018). Loss of time-dependent physiology associates, for example, with higher mortality in hypertensives with a non-dipping compared to a dipping blood pressure phenotype (Ben-Dov et al. 2007). Misalignment of the sleep-wake rhythms in healthy volunteers in a nightshift model increased blood pressure and inflammatory markers while suppressing heart rate variability (Morris et al. 2016) and blunted rhythmic gene expression profiles relevant for cellular immune response, apoptosis, cell proliferation and differentiation (Kervezee et al. 2018).

The molecular circadian clock is challenging to study under real-world conditions because this oscillator and its downstream rhythmic effects are modulated by environmental and behavioral components, that is, light exposure, sleep, food intake, and social interaction for the most parts. Examining biological rhythms in wrist temperature in the UK Biobank (Brooks et al. 2023), we argued that roughly 60% of the 6 °C fluctuations within 24 hours can be attributed to the circadian clock, while 25-30% are driven by behavioral and environmental factors. Since the latter factors are usually non-randomly distributed over the 24-hour cycle and can vary significantly between individuals, they introduce substantial measurement noise. To address this, we developed the human chronobiome study paradigm, where, despite these limitations, we can discern time-specific phenotypes through phenomic and multi-omic outputs in small cohorts of volunteers under real-world conditions (Skarke et al. 2017).

Data streams from wearable devices are increasingly used in clinical medicine for risk prediction and stratification (Spatz et al. 2024) and create opportunities for clinical phenotyping and determining disease risk. However, little is known about the prognostic value of out-of-clinic biometric monitoring of heart health in chronic kidney disease (CKD) (Wieringa et al. 2017). Under controlled clinical conditions, we observed a clear association between heart rate variability (HRV) and risk of mortality in QRS complexes over 10 seconds from a 12-lead resting electrocardiogram measured at baseline in 3,245 Chronic Renal Insufficiency Cohort (CRIC) participants with a median 4.2 years of follow-up (Drawz et al. 2013). A particular aspect we are interested in is to understand how the diurnal variability observed in time-series measurements relates to future disease risk. In the CRIC study, we found that rhythmic profiling of 24-hour blood pressure (BP) traces identified subgroups of CRIC participants at higher risk of cardiovascular death. CRIC participants with a history of cardiovascular disease (CVD) and absent cyclic components in their BP profile had at any time a 3.4-times higher risk of cardiovascular death than CVD patients with cyclic components present in their BP profile (HR: 3.38, 95% CI: 1.45-7.88, *p*=0.005) (Jamal et al. 2023).

Recently, we demonstrated feasibility for out-of-clinic evaluation of HRV in a two-center cross-sectional sample of CRIC participants and healthy controls (Lahens et al. 2022). We found preliminary evidence of deconsolidated rhythmic organization of HRV where, for example, the time-of-day dependent modulation of RR-intervals was lower in participants compared to controls. This work provided the roadmap for the current larger cross-sectional sample of CRIC participants. The goal of the current study was to confirm feasibility of obtaining HRV data in a large patient population, and to study the association between HRV and its time-specific variability associated with demographics, disease phenotypes, and therapeutic drug use. This adds to the growing body (Drawz et al. 2013; Lahens et al. 2022; Jamal et al. 2023) of predictive and time-specific phenotyping in CRIC participants using real-world sampling strategies.

## Results

The cross-sectional cohort consisted of 458 participants with valid BioPatch measurements collected from seven CRIC centers between 2019 and 2023. The study population had a mean age (and standard deviation) of 68.6±9.7 years, while 45% were female, 49.3% had diabetes, and 30.8% had pre-existing heart disease. Each participant contributed a mean (±SD) of 50.3±9.3 hours of BioPatch EKG recordings, including the heart rate variability (HRV) output SDNN (Standard Deviation of Normal-to-Normal intervals). SDNN is the standard deviation of the RR intervals under conditions of a sinus rhythm. To conduct a comparative analysis of how SDNN profiles are associated with disease risk, we split the multi-day data recordings of this cohort by SDNN tertiles into low SDNN (<33.7 ms; n=152), mid SDNN (33.7-48.8 ms; n=153), and high SDNN categories (>49.0 ms; n=153). For comparison, the normative reference value clinically established for short-term SDNN is 50±16 ms (Nunan, Sandercock, and Brodie 2010) and low SDNN values have been associated with increased cardiovascular risk (Kleiger et al. 1987). The distribution of age, sex, race, and ethnicity were similar between the tertiles (*p*>0.05, *Table 1*). This established comparability since we observed significant age- and sex-specific effects on SDNN. After multivariable adjustment, age increased SDNN by 0.22 ms/year (*p*=0.047, Table 2), and women had a 3.85 ms lower SDNN compared with men (*p*=0.06).

**Table 1.** Clinical and demographic characteristics of study population by SDNN tertiles.

| Table 1 |  | Tertile of SDNN |  |  |  |
| --- | --- | --- | --- | --- | --- |
| Characteristics | Overall (n=458) | Low (< 33.7) n=152 | Mid (33.8-48.8) n=153 | High (>49.0) n=153 | P value* |
| Age (years) | 68.6 (9.7) | 67.6 (9.1) | 68.2 (10.0) | 69.9 (10.0) | 0.11 |
| Sex |  |  |  |  |  |
| Female | 206 (45.0%) | 74 (48.7%) | 68 (44.4%) | 64 (41.8%) | 0.48 |
| Male | 252 (55.0%) | 78 (51.3%) | 85 (55.6%) | 89 (58.2%) |  |
| Race-Ethnicity |  |  |  |  |  |
| Hispanic | 75 (16.4%) | 28 (18.4%) | 31 (20.3%) | 16 (10.5%) | 0.1 |
| Non-Hispanic Black | 132 (28.8%) | 46 (30.3%) | 39 (25.5%) | 47 (30.7%) |  |
| Non-Hispanic White | 215 (46.9%) | 62 (40.8%) | 72 (47.1%) | 81 (52.9%) |  |
| Other | 36 (7.9%) | 16 (10.5%) | 11 (7.2%) | 9 (5.9%) |  |
| Diabetes | 226 (49.3%) | 100 (65.8%) | 70 (45.8%) | 56 (36.6%) | <0.0001 |
| History of Cardiovascular Disease | 141 (30.8%) | 53 (34.9%) | 33 (21.6%) | 55 (35.9%) | 0.01 |
| Hypertension | 411 (89.9%) | 140 (92.7%) | 142 (92.8%) | 129 (84.3%) | 0.02 |
| Diastolic BP (mmHg) | 70.0 (11.4) | 70.2 (12.1) | 72.1 (10.9) | 67.6 (10.9) | 0.003 |
| Systolic BP (mmHg) | 126.2 (17.2) | 126.4 (18.0) | 127.8 (17.1) | 124.4 (16.5) | 0.23 |
| Mean Arterial Pressure | 88.7 (11.8) | 89.0 (12.4) | 90.6 (11.5) | 86.5 (11.3) | 0.01 |
| Hemoglobin A1C (%) | 6.36 (1.34) | 6.85 (1.69) | 6.18 (0.98) | 6.05 (1.10) | <0.0001 |
| Urine PCR median (IQR) | 0.17 (0.07 - 0.60) | 0.27 (0.10 - 0.72) | 0.16 (0.07 - 0.48) | 0.14 (0.06 - 0.62) | 0.03 |
| Urine PCR [n (%)] |  |  |  |  |  |
| uPCR<0.2 | 250 (55.7%) | 68 (46.3%) | 90 (59.2%) | 92 (61.3%) | 0.02 |
| uPCR≥0.2 | 199 (44.3%) | 79 (53.7%) | 62 (40.8%) | 58 (38.7%) |  |
| eGFR (ml/min/1.73m<sup>2</sup>) | 51.1 (20.6) | 49.2 (20.4) | 52.3 (20.4) | 51.7 (21.1) | 0.37 |
| eGFR<45 | 182 (39.7%) | 64 (42.1%) | 57 (37.3%) | 61 (39.9%) | 0.69 |
| eGFR≥45 | 276 (60.3%) | 88 (57.9%) | 96 (62.7%) | 92 (60.1%) | . |
| BMI (kg/m<sup>2</sup>) | 31.2 (6.7) | 32.2 (6.3) | 31.1 (6.5) | 30.3 (7.2) | 0.05 |
| Smoking Status |  |  |  |  | 0.07 |
| Never Smoker | 260 (56.8%) | 78 (51.3%) | 98 (64.1%) | 84 (54.9%) |  |
| Smoker | 198 (43.2%) | 74 (48.7%) | 55 (35.9%) | 69 (45.1%) |  |
| Beta Blocker Use, n (%) | 184 (40.2%) | 60 (39.5%) | 65 (42.5%) | 59 (38.6%) | 0.8 |

N(%) for categorical variables; Mean(SD) for continuous variables
\* Chi-Square test for categorical variables; ANOVA for continuous variables.
uPCR: urine protein to creatinine ratio. It is unitless since both urine protein and creatine has unit of mg/dL.
eGFR: estimated glomerular filtration rate
BMI: body mass index

**Table 2.** Multivariable Linear Regression Results

| **Dependent Variable: SDNN in ms** |  |  |  |
| --- | --- | --- | --- |
| **Parameter** | **Estimate** | **SD** | **p value** |
| Age (years) | 0.22 | 0.11 | 0.047 |
| Sex Female vs Male | -3.85 | 2.04 | 0.06 |
| Diabetes Yes vs No | -7.41 | 2.10 | 0.001 |
| History of cardiovascular disease (CVD) Yes vs No | 1.13 | 2.26 | 0.62 |
| Hypertension Yes vs No | -6.18 | 3.57 | 0.084 |
| Smoker Yes vs No | 1.66 | 2.02 | 0.41 |
| Body Mass Index (BMI) | -0.072 | 0.16 | 0.65 |
| Urine Protein-to-Creatinine Ratio (uPCR) $\geq 0.2$ vs $< 0.2$ | 5.73 | 2.23 | 0.011 |
| Estimated Glomerular Filtration Rate (eGFR) $< 45$ vs $\geq 45$ | 5.32 | 2.20 | 0.016 |
| Beta Blocker Use Yes vs No | 1.87 | 2.23 | 0.40 |
| **Dependent Variable: SDNN - Amplitude in ms** |  |  |  |
| Age (years) | 0.074 | 0.043 | 0.083 |
| Sex Female vs Male | -2.23 | 0.78 | 0.005 |
| Diabetes Yes vs No | -1.45 | 0.81 | 0.074 |
| History of CVD Yes vs No | 0.16 | 0.87 | 0.86 |
| Hypertension Yes vs No | 0.92 | 1.37 | 0.51 |
| Smoker Yes vs No | 0.07 | 0.78 | 0.93 |
| BMI | 0.092 | 0.061 | 0.13 |
| uPCR $\geq 0.2$ vs $< 0.2$ | 0.16 | 0.86 | 0.85 |
| eGFR $< 45$ vs $\geq 45$ | -0.32 | 0.85 | 0.70 |
| Beta Blocker Use Yes vs No | -0.58 | 0.85 | 0.50 |
| **Dependent Variable: SDNN - Acrophase in hour** |  |  |  |
| Age (years) | 0.005 | 0.029 | 0.85 |
| Sex Female vs Male | 0.38 | 0.53 | 0.47 |
| Diabetes Yes vs No | 1.01 | 0.54 | 0.063 |
| History of CVD Yes vs No | 0.84 | 0.58 | 0.15 |
| Hypertension Yes vs No | -0.17 | 0.92 | 0.85 |
| Smoker Yes vs No | 0.02 | 0.52 | 0.96 |
| BMI* | -0.11 | 0.041 | 0.007 |
| uPCR $\geq 0.2$ vs $< 0.2$ | -0.13 | 0.58 | 0.83 |
| eGFR $< 45$ vs $\geq 45$ | 0.40 | 0.57 | 0.49 |
| Beta Blocker Use Yes vs No | 1.36 | 0.57 | 0.018 |

\* The estimate of -0.11 translates to a phase advance (left shift on the 24-hour scale of the x-axis) of SDNN peak levels by 6.6 minutes per unit of BMI increasing.

### Disease burden manifests in low heart rate variability

About half of the CRIC cohort had diabetes. A total of 65.8% of the participants in the low SDNN category were diabetic, compared to only 36.6% in the high SDNN category (*Table 1*). The burden of diabetes was evident in the multivariable linear regression where the presence of diabetes was significantly associated with a 7.4 ms lower SDNN compared to non-diabetic participants (*p*=0.001, *Table 2*). The association between diabetes and low levels of SDNN was recapitulated by a significant signal in the clinical laboratory assessment. Hemoglobin A1C levels were highest (6.85±1.69%) in the lowest SDNN tertile, and lower (6.18±0.98% and 6.05±1.10%, respectively) in the mid- and high-SDNN tertiles (*p*<0.0001, *Figure 1*, *Table 1*).

**Figure 1.**
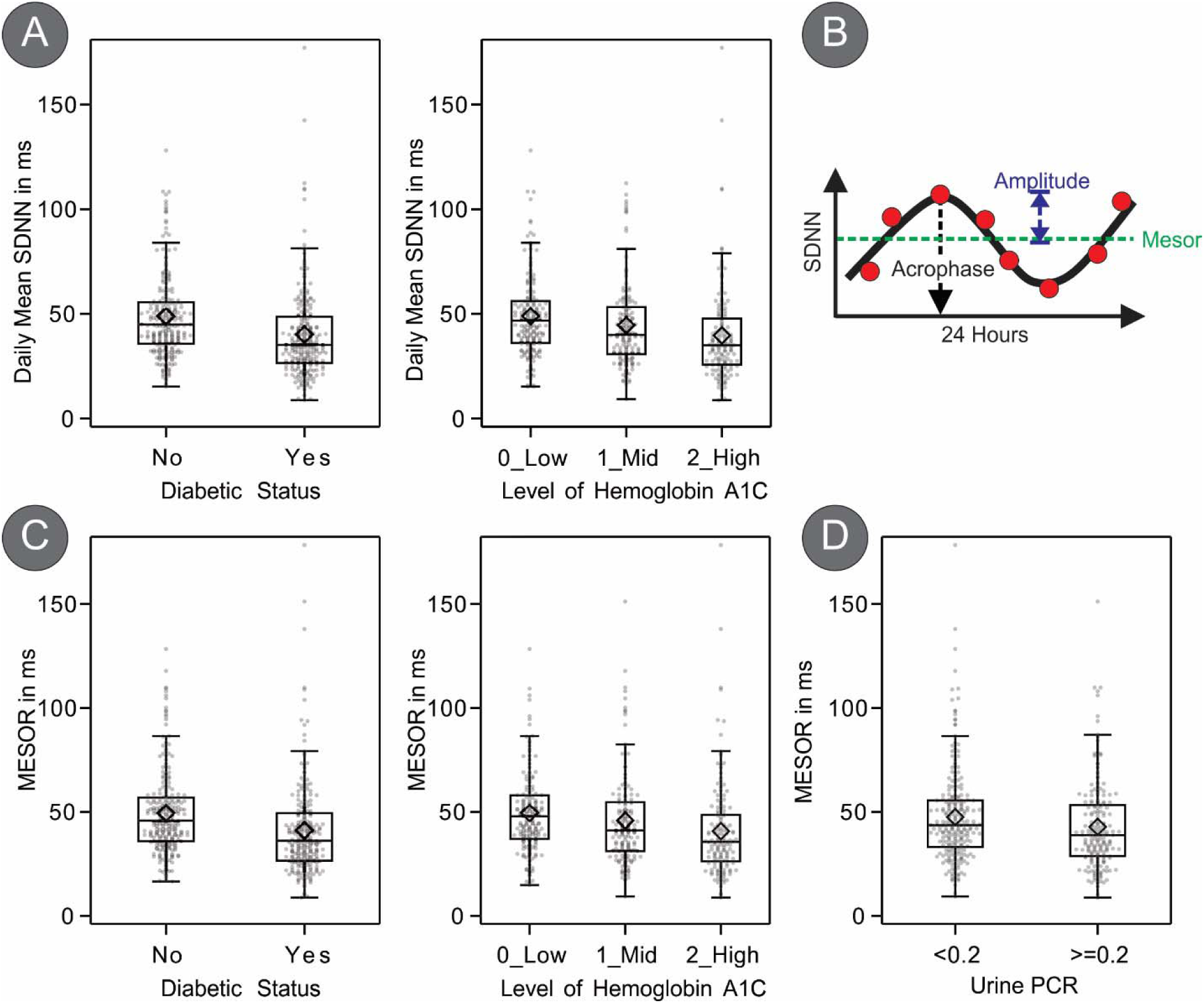
(A) Boxplots of daily mean SDNN by diabetic status (left) and tertiles of hemoglobin A1C levels (right) defined as low (4.5 to 5.6), mid (5.7 to 6.4), and high (6.5 to 14.4). (B) Schematic plot to show the outputs MESOR, amplitude, and phase from the cosinor statistical analysis. MESOR represents the midline-estimating statistic of rhythm. (C) Boxplots of SDNN MESOR by diabetic status (left) and tertiles of hemoglobin A1C levels (right). (D) Boxplot of SDNN MESOR for patients with high (uPCR≥0.2) versus low (uPCR<0.2) proteinuria level.

About a third of CRIC participants (n=141, 30.8%) reported a history of cardiovascular disease (CVD) which included heart failure, myocardial infarction, stroke, and peripheral artery disease (*p*=0.01, *Table 1*). Fewer participants with a history of CVD were present in the mid-SDNN tertile (n= 33, 21.6%) compared to the low (53, 34.9%) or high (55, 35.9%) SDNN tertiles (*p*= 0.01, *Table 1*), however, the multivariable linear regression analysis failed to reach significance (*p*=0.62, *Table 2*).

A significant trend (*p*=0.02, *Table 1*) was evident in the urine protein-creatinine-ratio (uPCR) across the SDNN tertiles. 53.7% of participants in the low SDNN tertile had high urine protein excretion (uPCR≥0.2) compared to 38.7% in the high SDNN tertile. Other parameters for renal function like eGFR did not show significant trends (*p*=0.37). In the multivariable linear regression, a significant interaction between uPCR and eGFR (*p value for test of interaction*=0.023) was noted where participants with compromised kidney function and proteinuria (eGFR<45 mL/min/1.73m^2^, uPCR≥0.2) showed a substantially lower SDNN of 41.9 ms compared to 53.6 ms in participants with compromised renal function but no signs of proteinuria (eGFR<45 mL/min/1.73m^2^, uPCR<0.2, *Table S 3*). This is reflective of how the presence of proteinuria in CKD potentiates the risk for CVD and underscores the emergence of proteinuria as an independent risk factor for cardiovascular morbidity and mortality (Currie and Delles 2013). Of note is that this effect was less pronounced in participants with a kidney function ≥45 mL/min/1.73m^2^ (SDNN of 41.9 ms in participants with proteinuria compared to 43.8 ms in participants without protein leakage).

### Disease-specific cyclical patterns in HRV

The rhythm-adjusted mean of SDNN (derived as MESOR [midline-estimating statistic of rhythm] from fitting a cosine function, *Figure 1B*) recapitulated the SDNN reported above (calculated as arithmetic means) (*Figure 1C,D)*. CRIC study participants with diabetes, for example, had lower SDNN MESOR compared with CRIC study participants without diabetes (41.2 ms vs. 49.5 ms, *p*<0.0001) and evident in the multivariable linear regression (a 7.4 ms decreased SDNN MESOR in participants with diabetes, *p*=0.0008, *Table 2*). SDNN MESORs tracked significantly with hemoglobin A1C plasma levels (*p*<0.0001). For proteinuria, participants with uPCR≥0.2 had lower SDNN MESOR compared to those with uPCR<0.2 (42.8 ms vs. 47.5 ms, *p*=0.0234) (*Figure 1D*). Multivariable linear regression results supported this observation; participants without proteinuria have a 5.15 ms higher SDNN compared to participants with proteinuria (*p*=0.027, *Table 2*). The elevation of SDNN levels during night hours was conserved independent of degree of proteinuria (*Figure 2B*).

**Figure 2.**
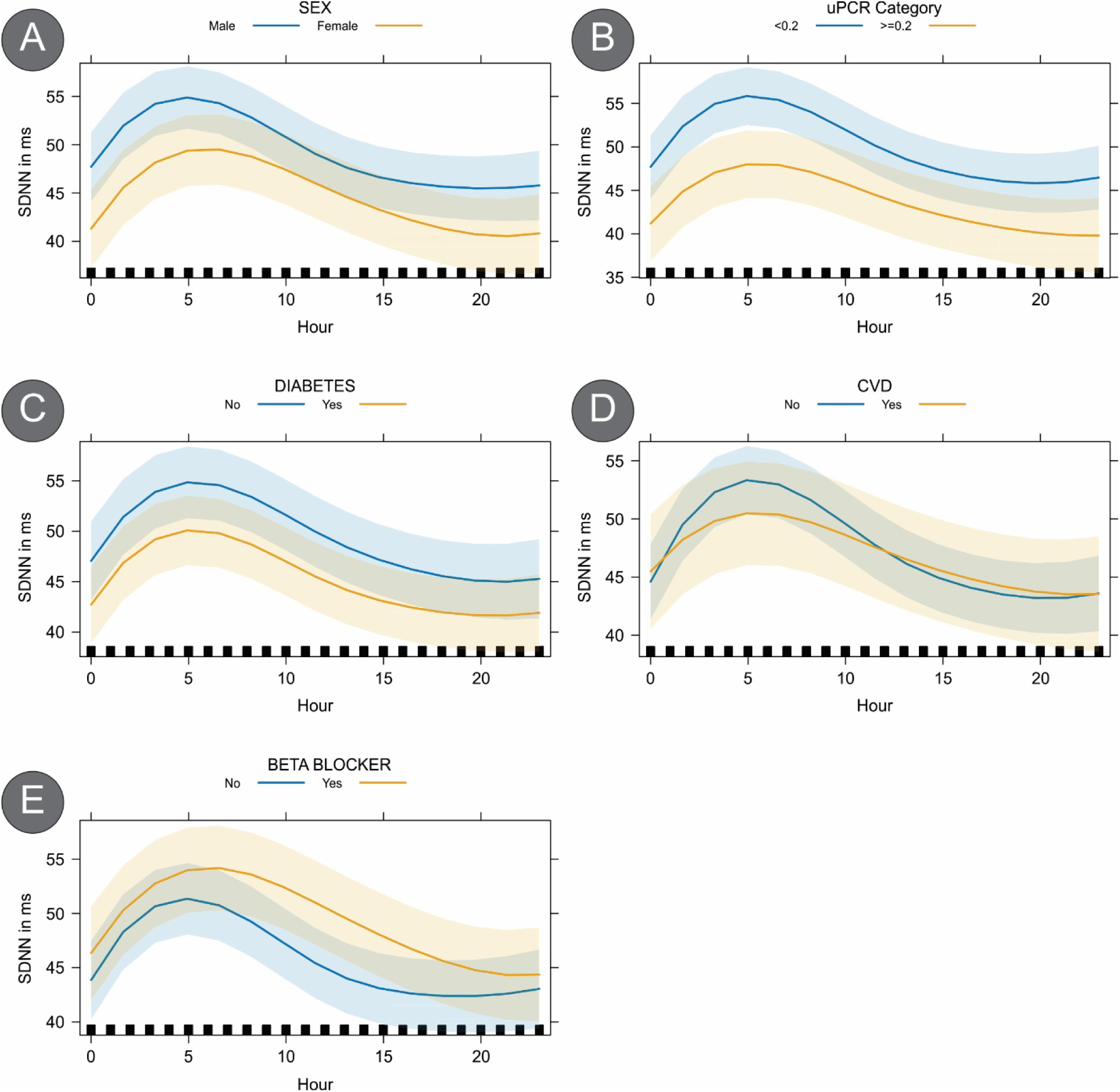
Spline-based estimation of the SDNN trace over 24 hours to characterize the differences in shapes of the curves for (A) male and female participants, (B) participants with different proteinuria levels, (C) diabetes diagnoses, (D) cardiovascular disease (CVD), (E) or beta blocker use.

We also divided participants into tertiles based on their SDNN amplitudes (Low: <5.1, n=152; Mid: 5.1-10.6, n=153; High: >10.6, n=153), quantified by the cosinor model (*Figure 1B; Table S 1b*), and observed significant differences in the proportion of participants with diabetes across the tertiles (*p*=0.006, *Table S 1b*). This corresponded to a 1.45 ms decrease in SDNN amplitude for participants with diabetes compared to those without in the multivariable linear regression model (*p*=0.074, *Table 2*). The SDNN amplitude tertile distribution of hemoglobin A1C did not attain statistical significance (*p*=0.26, *Table S 1b*).

Notably, men were over-represented in the high-SDNN amplitude tertile compared to the low- and medium-SDNN amplitude tertiles (*p*=0.047, *Table S 1b*). Since sex-specific modulation of short-term SDNN has been documented (Nunan, Sandercock, and Brodie 2010), we used splines to identify differences in the shape of curves. We found that men compared to women displayed higher SDNN values over the 24-hour time frame (*p*<.0001, *Figure 2A*) which replicates known physiology (Nunan, Sandercock, and Brodie 2010). Our multivariable linear regression estimated this sex-specific difference as a 2.23 ms higher SDNN amplitude in males compared to females (*p*=0.005, *Table 2*).

The last parameter we examined as output from the cosinor model was acrophase (*Table S 1c*). This parameter delineates where on the 24-hour clock time the peak of a sinusoidal curve lands. Heart rate variability outputs typically reach peak levels throughout the early morning hours as described by our group (Lahens et al. 2022) and others (Yoshizaki et al. 2013). To facilitate interpretation, we divided the CRIC cohort into four categories based on SDNN acrophase values: early morning (0AM-6AM, n=159), morning (6AM-12PM, n=188), afternoon (12PM-6PM, n=74) and evening hours (6PM-12AM, n=37). Note that acrophase is a circular parameter where a peak late in the evening hours is closely related to a peak in the early morning. We found significant trends for sex-specific differences between these SDNN acrophase quartiles (*p*=0.03), for diabetes (*p*=0.02), history of cardiovascular disease (*p*=0.003), eGFR (*p*=0.04), systolic blood pressure (*p*=0.04), and for beta blocker use (*p*=0.0002). In the spline analysis, curve shape differences supportive of acrophase shifts were evident for sex (*p*<0.0001, *Figure 2A* as described earlier). For participants with diabetes, the oscillatory SDNN curve was comparable with non-diabetic participants (**Error! Reference source not found.***C*). And for participants with cardiovascular disease the curve characteristics were significantly different between cases and controls (*p*=0.0005, **Error! Reference source not found.***D*) but timing of peak levels of SDNN, the acrophase, still occurred at a similar time of day. Furthermore, the spline analysis confirmed that the SDNN-over-time curve was significantly different between users and non-users of beta blockers (*p*=0.0005, **Error! Reference source not found.***E*). CRIC participants reporting beta blocker use showed an overall higher SDNN compared to non-users. This is in line with beta-blocker-induced increase of parasympathetic activity, resultant in increased SDNN (Niemela, Airaksinen, and Huikuri 1994).

The multivariable linear regression modeling allowed us to inspect phase shifts at a more granular level. Both diabetes diagnosis and beta blocker use were suggestive of phase delays in peak SDDN levels among the CRIC study participants (diabetes, 1 hour delay, *p*=0.06; beta blocker use, 1.36 hour delay, *p*=0.02; *Table 2*). The signal detected for BMI suggests that SDNN peak levels were phase advanced by 6.6 minutes per unit of BMI increase (*p*=0.007, *Table 2*). Though this relationship could be confounded by age (and age’s association with earlier chronotypes (Fischer et al. 2017)), indication, and other biases, this finding could suggest an altered cardiac autonomic state and thus may introduce a rhythm-based risk factor for CVD (Yadav et al. 2017).

## Discussion

This study assembled the largest dataset to date of SDNN as an index for heart rate variability from out-of-clinic wearable devices in the Chronic Renal Insufficiency Cohort (CRIC). The cohort structure allowed us to confirm sex- and disease-specific modulation of heart rate variability, most notably for diabetes and HbA1C levels as well as for kidney damage (uPCR). Differences in the time-specific variance of heart rate variability were evident for sex, disease and therapeutic drug use, where beta blocker use was associated with elevated heart rate variability. The value of the present study is that this sets the stage to explore in this CRIC population the predictiveness of HRV metrics for future CVD events and other future clinical outcomes.

The strengths of this study design include that the BioPatch devices were worn by CRIC participants during their activities of daily living, that participants wore these devices for at least two complete circadian cycles (over 48 hours) to enhance detection and credibility of rhythmic features (Hughes et al. 2017), and that data were collected in a multi-center effort. These characteristics strengthen the applicability of our findings to the general CKD population and beyond.

SDNN values in the present study are consistent with normative data developed clinically from short-term EKG recordings (Nunan, Sandercock, and Brodie 2010) and align well with our preliminary results based on 48-hour recordings (Lahens et al. 2022), though at much higher precision. The heterogeneity of the study cohort allowed us to determine associations of SDNN summary and rhythmic readouts with demographic factors, renal function and comorbidities. Here, as expected, the significantly lower SDNN in CRIC participants with diabetes stood out as a risk factor. Known is that annual mortality rates increase dramatically as SDNN decreases. This is described in a similarly powered study in chronic heart failure from the UK using 24-hour ambulatory EKGs determined risk categories with the mortality rising from 5.5% for patients with SDNN>100 ms to 12.7% for SDNN between 50 and 100 ms, up to 51.4% for SDNN<50 ms (Nolan et al. 1998). The mean of SDNN 50.3±9.3 in the present study is situated right at the threshold where the upward relationship between lower SDNN and increased risk of mortality (13%) transitions into the much steeper association of very low SDNN and high mortality risk (51%). In the context of the present study, the ∼4 ms the SDNN was significantly lower in CRIC participants with diabetes potentially exposes them to a disproportionately higher risk of mortality.

The shape of heart rate variability over the course of 24 hours allows for the time-specific characterization of disease risk phenotypes. Our data highlights, for example, that in CRIC participants with diabetes SDNN levels were lower over the entire 24-hour day compared to CRIC participants without diabetes. In contrast, participants with CVD showed reduced SDNN levels primarily during the nighttime hours compared to participants without CVD. This time-specific phenomenon has been associated with increased risk for stroke (Binici et al. 2011) and cardiovascular events (Hadad et al. 2021). This sets the stage for future studies in this CRIC subpopulation to parse how time-specific features of heart rate variability associate with renal function, CVD events, and mortality, which are not yet feasible due to limited follow-up time. Notably, we established the framework to address this approach in the UK Biobank where we were able to predict the future onset of 73 diseases by discerning rhythmic features in wrist body temperature traces collected with wearable fitness trackers under the noisy conditions of daily life (Brooks et al. 2023).

Limitations for the present study include that this is a correlative study with variables limited to demographic factors, comorbidity, and renal function. The short follow up period of just two years for some of the participants does not yet allow association studies between HRV parameters and clinical outcomes.

In conclusion, we established a paradigm to leverage wearable digital health technology to monitor time-dependent cardiovascular health in participants with CKD. This platform is essential to develop clinically actionable, digital biomarkers to improve management of CVD in CKD.

## Methods

The baseline data was acquired in CRIC participants from 2019-2020 (Phase 4). The design, inclusion and exclusion criteria, and baseline characteristics of the CRIC study participants have been published previously (Feldman et al. 2003; Lash et al. 2009). In brief, participants where included if they had not reached ESRD (end-stage renal disease) and consented to participation in the CRIC2018 (Phase IV), but excluded if they were a resident in a nursing home, non-ambulatory, equipped with heart pacemaker or automatic defibrillator, presented with a skin rash or irritation at the site of wearable placement, Modified Mini-Mental State Examination < 80, or ESRD. This study used the Zephyr BioPatch (Zephyr Technology, Annapolis, MD, USA), distributed by Medtronic Corporation (Minneapolis, MN, USA). This research-grade wearable device records cardiovascular, and respiratory activity. The target observational time for each CRIC participants was set to 48 hours to cover two consecutive 24-hour circadian cycles in order to enhance the detection of biological rhythms. Participants received a plug-in charging cradle and were instructed to remove the BioPatch device from its chest-mounted holder after approximately 24 hours of use to recharge the device for approximately two hours followed by inserting it back into the BioPatch holder on the chest for continued recordings. Positioning of the two EKG snap electrodes to hold the BioPatch holder in place followed standard EKG guidelines for V1 and V2, i.e. V1 corresponds to the right fourth intercostal space; and V2 corresponds to the left fourth intercostal space. Prior to application of the snap electrodes skin was cleaned with rubbing alcohol and shaved if necessary.

We first divided the raw data into hourly intervals. Within each one-hour interval, we calculated the average indices of SDNN as the metric of HRV for this study. SDNN was defined as the Standard Deviation of Normal-to-Normal (NN) Intervals measured in the ECG trace from normal R-R intervals in a rolling window of 300 beats (∼5 min). Quality control led to the exclusion of n=125 participant recordings. A majority of those (n=86) had episodes of ≥5-hour consecutive missingness of HRV traces (*Figure S 1*) which compromised confidence in the analysis of diurnal heart rate variability.

Using the summary data from each hourly interval, we replicated analyses published in our CRIC BioPatch pilot study (Lahens, Rahman et al. 2022). In sensitivity analyses, we varied the length of time interval (e.g., 30-min, 10-min) to evaluate how sensitive the analysis is with respect to the time interval and found similar results.

We modeled the SDNN using two approaches. Our first approach was to implement cosinor analysis for rhythm detection (Cornelissen 2014) to quantify MESOR (midline-estimating statistic of rhythm), amplitude, and acrophase (Figure 2) for each individual. Note, that since CRIC participants are monitored in the outpatient environment and entrained to a 24-hour rhythm, period in this model is fixed at 24 hours. We then fit separate linear regression models to identify risk factors that are associated with SDNN MESOR, amplitude, and phase. Variable selection was done through backward elimination with p-value threshold of 0.1. A covariate that was retained in any of the three models after variable selection was included in all three final models. The final models included the following covariates: age, sex, race/ethnicity, diabetic status, hypertension, history of cardiovascular disease, smoker, BMI, eGFR, uPCR, and beta blocker use. The second approach was to fit linear mixed effects model on hourly SDNN data directly. In the model, we used restricted cubic splines to allow flexible HRV pattern over time within the day. Here, the advantage is that the modeling process captures the exact shape of the diurnal variability. The model included a random intercept at an individual level and adjusted for the same covariates as in the first approach.

## Supplementary Figures

**Figure S 1.**
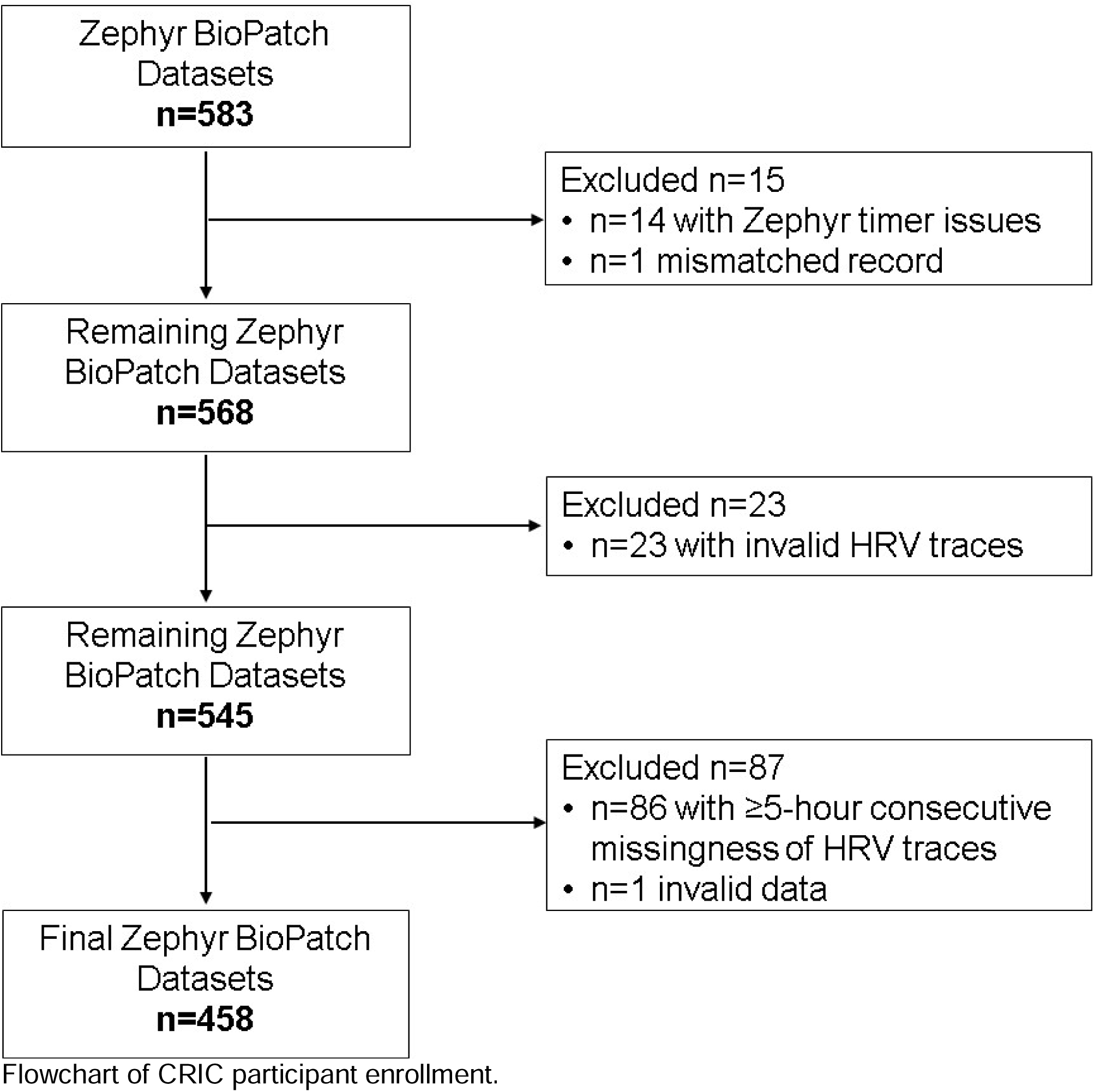
Flowchart of CRIC participant enrollment.

## Supplementary Tables

**Table S 1.**
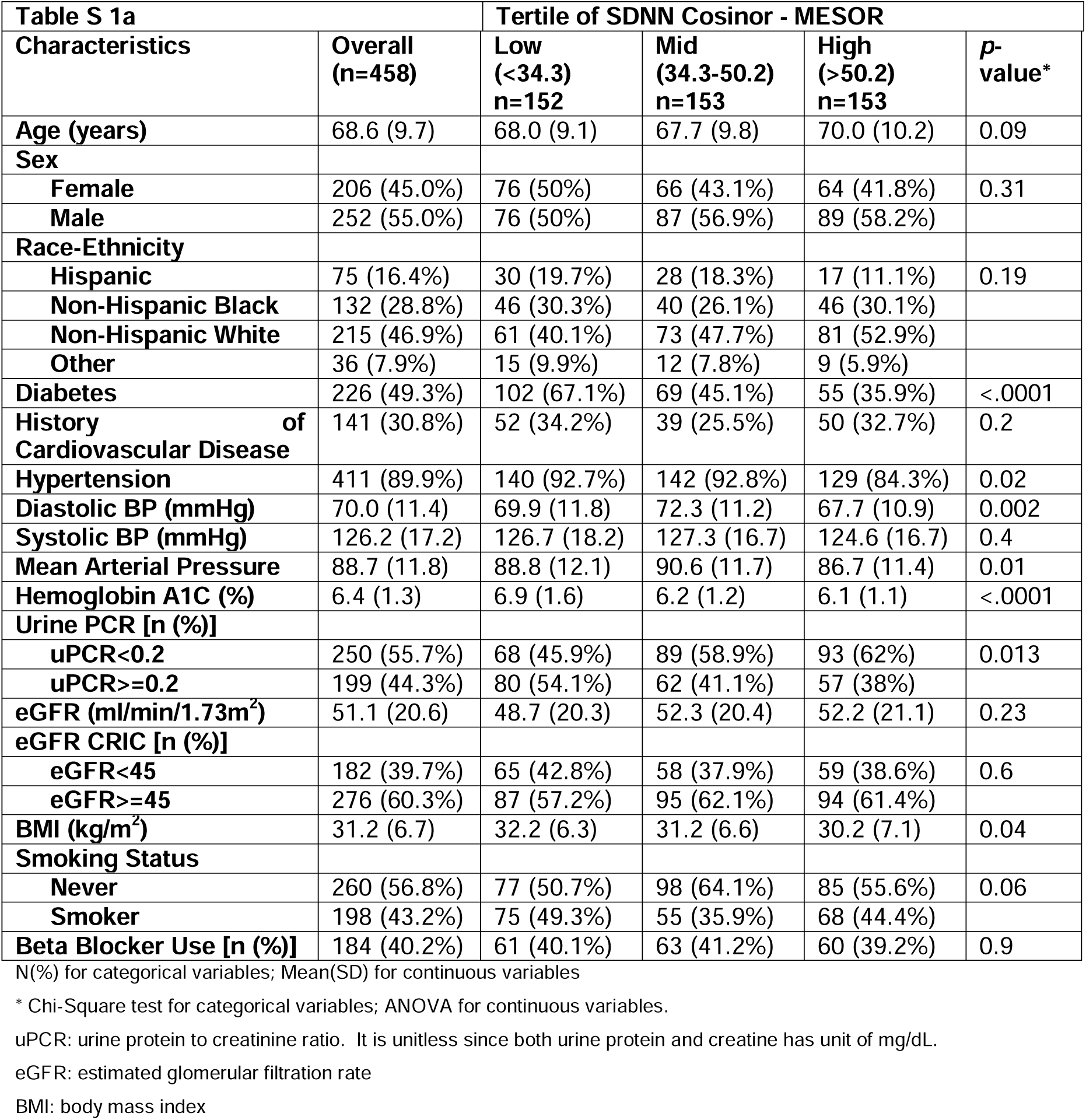
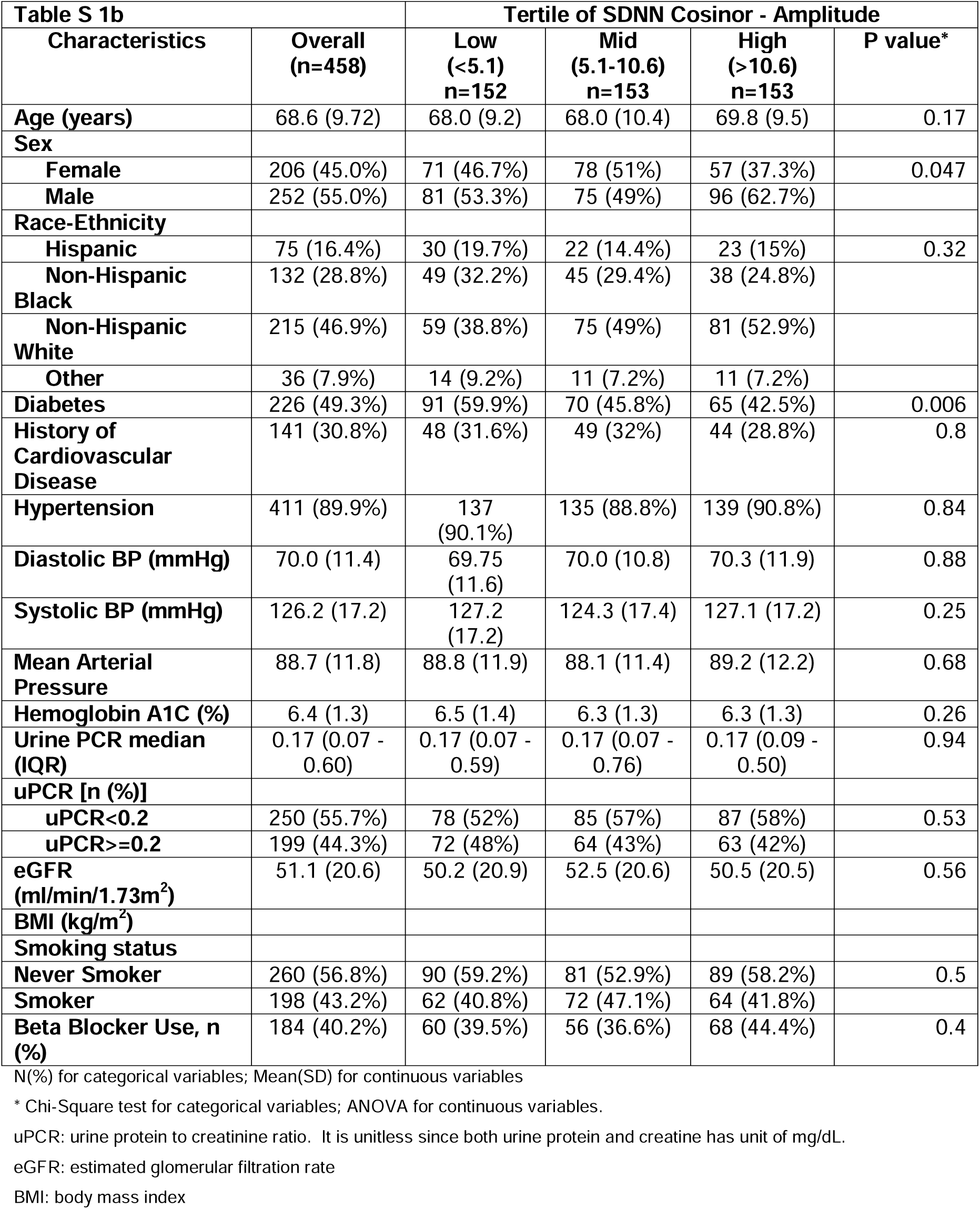
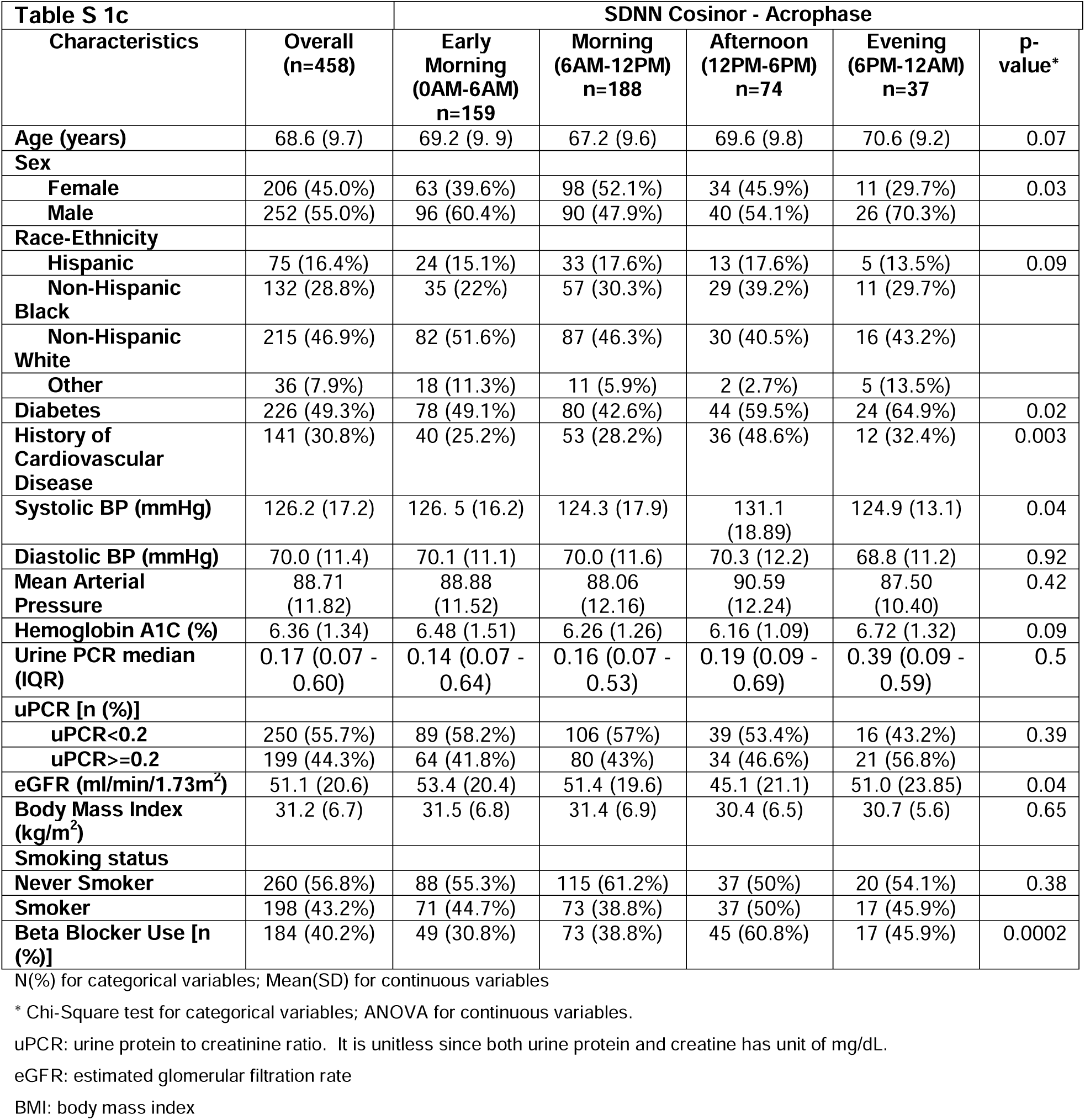
Clinical and demographic characteristics of study population by SDNN MESOR, Amplitude, or Acrophase tertiles.

**Table S 2.** Multivariate Linear Regression Results SDNN MESOR

| **Dependent Variable: SDNN - MESOR in ms** |  |  |  |
| --- | --- | --- | --- |
| Parameter | Estimate | SD | p-value |
| Age (years) | 0.26 | 0.12 | 0.028 |
| Sex Female vs Male | -4.37 | 2.13 | 0.041 |
| Diabetes Yes vs No | -7.4 | 2.19 | 0.0008 |
| History of CVD Yes vs No | 0.83 | 2.35 | 0.73 |
| Hypertension Yes vs No | -6.7 | 3.72 | 0.073 |
| Smoker Yes vs No | 1.34 | 2.1 | 0.53 |
| BMI | -0.02 | 0.17 | 0.93 |
| uPCR $\geq 0.2$ vs $< 0.2$ | 5.15 | 2.33 | 0.027 |
| eGFR $< 45$ vs $\geq 45$ | 4.95 | 2.3 | 0.032 |
| Beta Blocker Use Yes vs No | 2.1 | 2.32 | 0.37 |

**Table S 3.** Estimated SDNN accounting for the Interaction between eGFR and uPCR

| **SDNN [ms]** | **eGFR$\geq 45$** | **eGFR$< 45$** |
| --- | --- | --- |
| **PCR$< 0.2$** | 43.8 | 53.6 |
| **PCR$\geq 0.2$** | 41.9 | 41.9 |

## Data Availability

All data produced in the present study are available upon reasonable request from the CRIC Study Investigators.

## Acknowledgments

We are indebted to the CRIC participants for volunteering in this study. Funding for the CRIC Study was obtained under a cooperative agreement from National Institute of Diabetes and Digestive and Kidney Diseases (U01DK060990, U01DK060984, U01DK061022, U01DK061021, U01DK061028, U01DK060980, U01DK060963, U01DK060902 and U24DK060990). In addition, this work was supported in part by: the Perelman School of Medicine at the University of Pennsylvania Clinical and Translational Science Award NIH/NCATS UL1TR000003, Johns Hopkins University UL1 TR-000424, University of Maryland GCRC M01 RR-16500, Clinical and Translational Science Collaborative of Cleveland, UL1TR000439 from the National Center for Advancing Translational Sciences (NCATS) component of the National Institutes of Health and NIH roadmap for Medical Research, Michigan Institute for Clinical and Health Research (MICHR) UL1TR000433, University of Illinois at Chicago CTSA UL1RR029879, Tulane COBRE for Clinical and Translational Research in Cardiometabolic Diseases P20 GM109036, Kaiser Permanente NIH/NCRR UCSF-CTSI UL1 RR-024131, Department of Internal Medicine, University of New Mexico School of Medicine Albuquerque, NM R01DK119199. CS is the Robert L McNeil Jr. Endowed Fellow in Translational Medicine and Therapeutics.

## References

1. Ben-Dov, I. Z., J. D. Kark, D. Ben-Ishay, J. Mekler, L. Ben-Arie, and M. Bursztyn. 2007. ’Predictors of all-cause mortality in clinical ambulatory monitoring: unique aspects of blood pressure during sleep’, Hypertension, 49: 1235–41.

2. Binici, Z., M. R. Mouridsen, L. Kober, and A. Sajadieh. 2011. ’Decreased nighttime heart rate variability is associated with increased stroke risk’, Stroke, 42: 3196–201.

3. Brooks, T. G., N. F. Lahens, G. R. Grant, Y. I. Sheline, G. A. FitzGerald, and C. Skarke. 2023. ’Diurnal rhythms of wrist temperature are associated with future disease risk in the UK Biobank’, Nat Commun, 14: 5172.

4. Cornelissen, G. 2014. ’Cosinor-based rhythmometry’, *Theor Biol Med Model*, 11: 16. Currie, G., and C. Delles. 2013. ’Proteinuria and its relation to cardiovascular disease’, Int J Nephrol Renovasc Dis, 7: 13–24.

5. Drawz, P. E., D. C. Babineau, C. Brecklin, J. He, R. R. Kallem, E. Z. Soliman, D. Xie, D. Appleby, A. H. Anderson, M. Rahman, and Cric Study Investigators. 2013. ’Heart rate variability is a predictor of mortality in chronic kidney disease: a report from the CRIC Study’, Am J Nephrol, 38: 517–28.

6. Feldman, H. I., L. J. Appel, G. M. Chertow, D. Cifelli, B. Cizman, J. Daugirdas, J. C. Fink, E. D. Franklin-Becker, A. S. Go, L. L. Hamm, J. He, T. Hostetter, C. Y. Hsu, K. Jamerson, M. Joffe, J. W. Kusek, J. R. Landis, J. P. Lash, E. R. Miller, E. R. Mohler, 3rd, P. Muntner, A. O. Ojo, M. Rahman, R. R. Townsend, J. T. Wright, and Investigators Chronic Renal Insufficiency Cohort Study. 2003. ’The Chronic Renal Insufficiency Cohort (CRIC) Study: Design and Methods’, J Am Soc Nephrol, 14: S148–53.

7. Fischer, D., D. A. Lombardi, H. Marucci-Wellman, and T. Roenneberg. 2017. ’Chronotypes in the US - Influence of age and sex’, PLoS One, 12: e0178782.

8. Hadad, R., B. S. Larsen, P. Weber, D. Stavnem, O. P. Kristiansen, O. W. Nielsen, S. B. Haugaard, and A. Sajadieh. 2021. ’Night-time heart rate variability identifies high-risk people among people with uncomplicated type 2 diabetes mellitus’, Diabet Med, 38: e14559.

9. Hughes, M. E., K. C. Abruzzi, R. Allada, R. Anafi, A. B. Arpat, G. Asher, P. Baldi, C. de Bekker, D. Bell-Pedersen, J. Blau, S. Brown, M. F. Ceriani, Z. Chen, J. C. Chiu, J. Cox, A. M. Crowell, J. P. DeBruyne, D. J. Dijk, L. DiTacchio, F. J. Doyle, G. E. Duffield, J. C. Dunlap, K. Eckel-Mahan, K. A. Esser, G. A. FitzGerald, D. B. Forger, L. J. Francey, Y. H. Fu, F. Gachon, D. Gatfield, P. de Goede, S. S. Golden, C. Green, J. Harer, S. Harmer, J. Haspel, M. H. Hastings, H. Herzel, E. D. Herzog, C. Hoffmann, C. Hong, J. J. Hughey, J. M. Hurley, H. O. de la Iglesia, C. Johnson, S. A. Kay, N. Koike, K. Kornacker, A. Kramer, K. Lamia, T. Leise, S. A. Lewis, J. Li, X. Li, A. C. Liu, J. J. Loros, T. A. Martino, J. S. Menet, M. Merrow, A. J. Millar, T. Mockler, F. Naef, E. Nagoshi, M. N. Nitabach, M. Olmedo, D. A. Nusinow, L. J. Ptacek, D. Rand, A. B. Reddy, M. S. Robles, T. Roenneberg, M. Rosbash, M. D. Ruben, S. S. C. Rund, A. Sancar, P. Sassone-Corsi, A. Sehgal, S. Sherrill-Mix, D. J. Skene, K. F. Storch, J. S. Takahashi, H. R. Ueda, H. Wang, C. Weitz, P. O. Westermark, H. Wijnen, Y. Xu, G. Wu, S. H. Yoo, M. Young, E. E. Zhang, T. Zielinski, and J. B. Hogenesch. 2017. ’Guidelines for Genome-Scale Analysis of Biological Rhythms’, J Biol Rhythms, 32: 380–93.

10. Jamal, N. E., T. G. Brooks, J. Cohen, R. R. Townsend, G. Rodriguez de Sosa, V. Shah, R. G. Nelson, P. E. Drawz, P. Rao, Z. Bhat, A. Chang, W. Yang, G. A. FitzGerald, C. Skarke, and Cric Study Investigators. 2023. ’Prognostic utility of rhythmic components in 24-hour ambulatory blood pressure monitoring for the risk stratification of chronic kidney disease patients with cardiovascular co-morbidity’, *medRxiv*.

11. Juffre, A., and M. L. Gumz. 2024. ’Recent advances in understanding the kidney circadian clock mechanism’, Am J Physiol Renal Physiol, 326: F382–F93.

12. Kervezee, L., M. Cuesta, N. Cermakian, and D. B. Boivin. 2018. ’Simulated night shift work induces circadian misalignment of the human peripheral blood mononuclear cell transcriptome’, Proc Natl Acad Sci U S A, 115: 5540–45.

13. Kleiger, R. E., J. P. Miller, J. T. Bigger, Jr., and A. J. Moss. 1987. ’Decreased heart rate variability and its association with increased mortality after acute myocardial infarction’, Am J Cardiol, 59: 256–62.

14. Lahens, N. F., M. Rahman, J. B. Cohen, D. L. Cohen, J. Chen, M. R. Weir, H. I. Feldman, G. R. Grant, R. R. Townsend, C. Skarke, and A. T. C. Study Investigators. 2022. ’Time-specific associations of wearable sensor-based cardiovascular and behavioral readouts with disease phenotypes in the outpatient setting of the Chronic Renal Insufficiency Cohort’, Digit Health, 8: 20552076221107903.

15. Lash, J. P., A. S. Go, L. J. Appel, J. He, A. Ojo, M. Rahman, R. R. Townsend, D. Xie, D. Cifelli, J. Cohan, J. C. Fink, M. J. Fischer, C. Gadegbeku, L. L. Hamm, J. W. Kusek, J. R. Landis, A. Narva, N. Robinson, V. Teal, H. I. Feldman, and Group Chronic Renal Insufficiency Cohort Study. 2009. ’Chronic Renal Insufficiency Cohort (CRIC) Study: baseline characteristics and associations with kidney function’, Clin J Am Soc Nephrol, 4: 1302–11.

16. Morris, C. J., T. E. Purvis, K. Hu, and F. A. Scheer. 2016. ’Circadian misalignment increases cardiovascular disease risk factors in humans’, Proc Natl Acad Sci U S A, 113: E1402–11.

17. Niemela, M. J., K. E. Airaksinen, and H. V. Huikuri. 1994. ’Effect of beta-blockade on heart rate variability in patients with coronary artery disease’, J Am Coll Cardiol, 23: 1370–7.

18. Nolan, J., P. D. Batin, R. Andrews, S. J. Lindsay, P. Brooksby, M. Mullen, W. Baig, A. D. Flapan, A. Cowley, R. J. Prescott, J. M. Neilson, and K. A. Fox. 1998. ’Prospective study of heart rate variability and mortality in chronic heart failure: results of the United Kingdom heart failure evaluation and assessment of risk trial (UK-heart)’, Circulation, 98: 1510–6.

19. Nunan, D., G. R. Sandercock, and D. A. Brodie. 2010. ’A quantitative systematic review of normal values for short-term heart rate variability in healthy adults’, Pacing Clin Electrophysiol, 33: 1407–17.

20. Skarke, C., N. F. Lahens, S. D. Rhoades, A. Campbell, K. Bittinger, A. Bailey, C. Hoffmann, R. S. Olson, L. Chen, G. Yang, T. S. Price, J. H. Moore, F. D. Bushman, C. S. Greene, G. R. Grant, A. M. Weljie, and G. A. FitzGerald. 2017. ’A Pilot Characterization of the Human Chronobiome’, Sci Rep, 7: 17141.

21. Spatz, E. S., G. S. Ginsburg, J. S. Rumsfeld, and M. P. Turakhia. 2024. ’Wearable Digital Health Technologies for Monitoring in Cardiovascular Medicine’, The New England journal of medicine, 390: 346–56.

22. Thosar, S. S., M. P. Butler, and S. A. Shea. 2018. ’Role of the circadian system in cardiovascular disease’, J Clin Invest, 128: 2157–67.

23. Wieringa, F. P., N. J. H. Broers, J. P. Kooman, F. M. Van Der Sande, and C. Van Hoof. 2017. ’Wearable sensors: can they benefit patients with chronic kidney disease?’, Expert Rev Med Devices, 14: 505–19.

24. Yadav, R. L., P. K. Yadav, L. K. Yadav, K. Agrawal, S. K. Sah, and M. N. Islam. 2017. ’Association between obesity and heart rate variability indices: an intuition toward cardiac autonomic alteration - a risk of CVD’, Diabetes Metab Syndr Obes, 10: 57–64.

25. Yoshizaki, T., Y. Kawano, Y. Tada, A. Hida, T. Midorikawa, K. Hasegawa, T. Mitani, T. Komatsu, and F. Togo. 2013. ’Diurnal 24-hour rhythm in ambulatory heart rate variability during the day shift in rotating shift workers’, J Biol Rhythms, 28: 227–36.

